# Inferring Viral Transmission Pathways from Within-Host Variation

**DOI:** 10.1101/2023.10.14.23297039

**Authors:** Ivan O. A. Specht, Brittany A. Petros, Gage K. Moreno, Taylor Brock-Fisher, Lydia A. Krasilnikova, Mark Schifferli, Katherine Yang, Paul Cronan, Olivia Glennon, Stephen F. Schaffner, Daniel J. Park, Bronwyn L. MacInnis, Al Ozonoff, Ben Fry, Michael D. Mitzenmacher, Patrick Varilly, Pardis C. Sabeti

**Affiliations:** The Broad Institute of MIT and Harvard, Cambridge, MA 02142, USA; Harvard College, Faculty of Arts and Sciences, Harvard University, Cambridge, MA 02138, USA; Harvard-MIT Program in Health Sciences and Technology, Cambridge, MA 02139, USA; Harvard/MIT MD-PhD Program, Boston, MA 02115, USA; Systems, Synthetic, and Quantitative Biology PhD Program, Department of Systems Biology, Harvard Medical School, Boston, MA 02115, USA; Department of Organismic and Evolutionary Biology, Faculty of Arts and Sciences, Harvard University, Cambridge, MA 02138, USA; Department of Immunology and Infectious Diseases, Harvard T.H. Chan School of Public Health, Harvard University, Boston, MA 02115, USA; Fathom Information Design, Boston, MA 02114, USA; Massachusetts Consortium on Pathogen Readiness, Harvard Medical School, Harvard University, Boston, MA 02115, USA; Department of Computer Science, School of Engineering and Applied Sciences, Harvard University, Cambridge, MA 02138, USA; Howard Hughes Medical Institute, Chevy Chase, MD 20815, USA

## Abstract

Genome sequencing can offer critical insight into pathogen spread in viral outbreaks, but existing transmission inference methods use simplistic evolutionary models and only incorporate a portion of available genetic data. Here, we develop a robust evolutionary model for transmission reconstruction that tracks the genetic composition of within-host viral populations over time and the lineages transmitted between hosts. We confirm that our model reliably describes within-host variant frequencies in a dataset of 134,682 SARS-CoV-2 deep-sequenced genomes from Massachusetts, USA. We then demonstrate that our reconstruction approach infers transmissions more accurately than two leading methods on synthetic data, as well as in a controlled outbreak of bovine respiratory syncytial virus and an epidemiologically-investigated SARS-CoV-2 outbreak in South Africa. Finally, we apply our transmission reconstruction tool to 5,692 outbreaks among the 134,682 Massachusetts genomes. Our methods and results demonstrate the utility of within-host variation for transmission inference of SARS-CoV-2 and other pathogens, and provide an adaptable mathematical framework for tracking within-host evolution.

## MAIN

Advances in pathogen genomic sequencing have enhanced our ability to determine infectious disease transmission pathways from the accumulation of mutations over time [1–20]. Inferred transmission networks provide critical insight into how pathogens spread, quantifying key parameters such as the effective reproductive number. They also permit evaluation of the effectiveness of mitigation strategies, e.g., vaccination [16] or nonpharmaceutical interventions [21,22] on transmission rates.

Numerous algorithms for reconstructing outbreaks based on genomic and epidemiological data have been developed [1,4,9,12,14,19,20,23]. The majority of existing models use the number of single nucleotide variants (SNVs) between each pair of cases as the only genetic signal to inform transmission. While a lower SNV distance is indeed a stronger indicator of transmission than a higher one, this approach oversimplifies the true underlying biological processes in a way that loses information [6,17], as it overlooks the mechanism that give rise to genetic variants in the first place. Moreover, these methods rely only on consensus genomes, despite the fact deep-sequencing data can capture intrahost single nucleotide variants (iSNVs).

Some transmission inference studies do model within-host evolution explicitly and incorporate iSNVs by adapting existing methods in phylogenetics [8,10,13]. This approach, while much more reflective of the underlying biological process, requires both enormous computational power and a highly efficient algorithmic implementation to yield accurate results if implemented in a Bayesian context [10]. And while maximum-likelihood methods yield results more efficiently [8,13], they fail to capture the different probabilities associated with different transmission networks, as well as the distributions of key epidemic parameters such as the mutation rate and serial interval.

In this paper, we develop, validate, and apply a novel evolutionary model for outbreak reconstruction that tracks the composition of the viral population within each host over time (**Figure 1**). Under our model, iSNV frequencies follow an approximate power-law distribution, a result we confirm empirically using a public dataset of 134,682 deep-sequenced SARS-CoV-2 genomes from Massachusetts, USA. Extending our within-host evolutionary model to outbreak reconstruction, we demonstrate that our approach infers transmission events more accurately then two leading transmission reconstruction methods, using synthetic, experimental animal, and human outbreaks for validation. Finally, we apply our model to a dataset of 5,692 outbreak clusters among the 134,682 Massachusetts genomes, quantifying the prevalence of genetic signatures in transmission links. Our findings improve upon existing methods in outbreak reconstruction while underscoring the usefulness of deep-sequencing data in epidemiological investigations.

**Figure 1:**
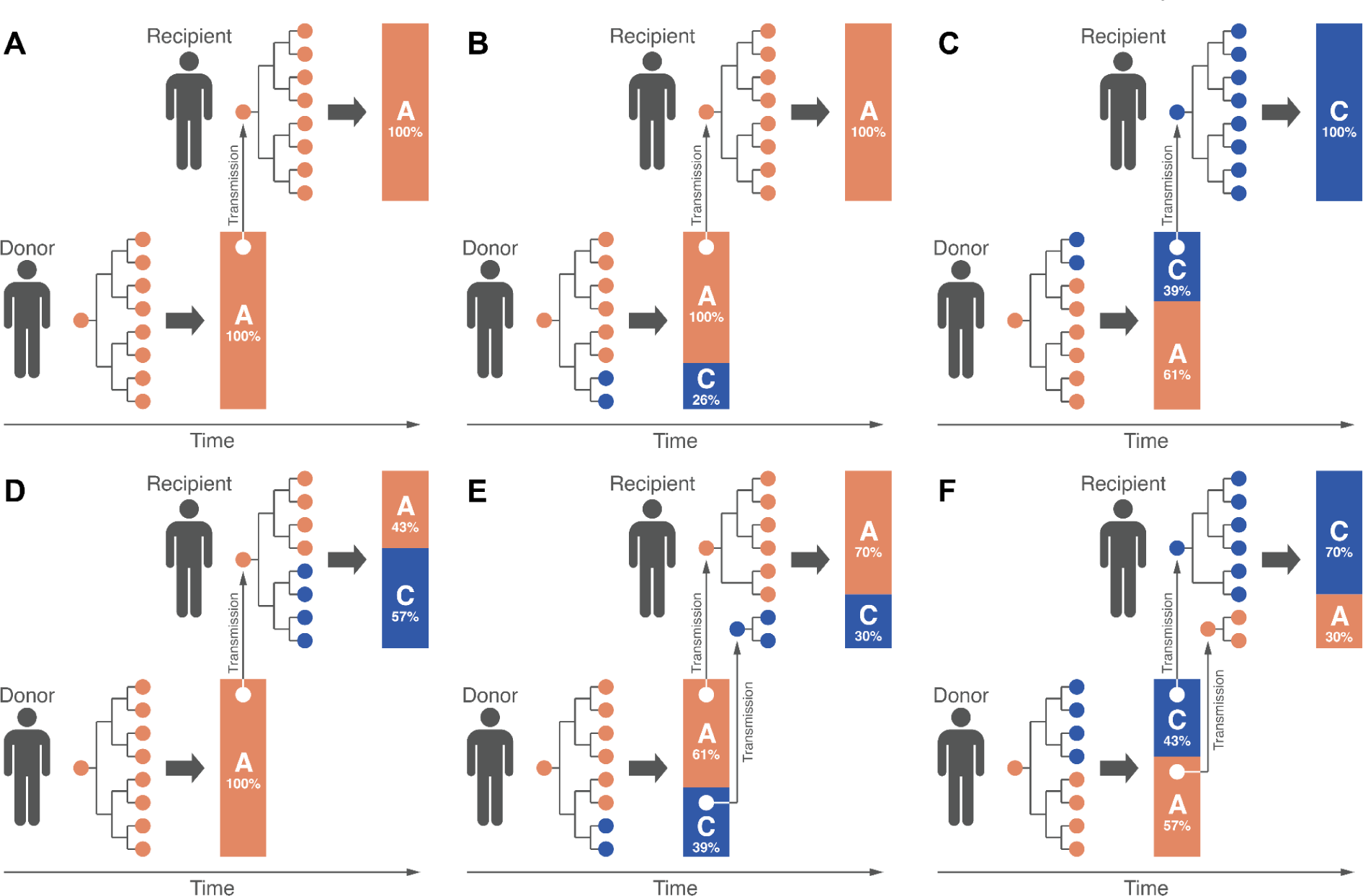
Schematic representations of six possible genetic profiles of viral transmission. In all diagrams, an orange circle represents a virion with adenine (A) at a given site *k* on the viral genome, and a blue circle represents a virion with cytosine (C) at site *k*. The branching process diagrams represent the early phase of infection (labeled Phase 1), when the viral population remains small, and mutation events, although rare, significantly impact the resulting minor allele frequency. The bar charts represent the proportion of virions exhibiting the major and minor allele at site *k* at the end of Phase 2, in which the viral population grows to its maximum size of approximately 10^9^ to 10^11^ particles [24]. The vertical gray arrows represent the virions passed from donor to recipient through the transmission bottleneck. In **(A)**, no iSNVs are observed at site *k* in the donor or recipient, with both exhibiting the same allele. In **(B)**, an iSNV arises at site *k* in the donor, and the major allele is passed to the recipient through a bottleneck of one particle. In **(C)**, an iSNV arises at site *k* in the donor, and the minor allele is passed to the recipient through a bottleneck of one particle, resulting in a change of consensus genome. In **(D)**, an iSNV arises at site *k* in the recipient, where it rises to consensus frequency, resulting in a change of consensus genome. In **(E)**, an iSNV arises at site *k* in the donor, and both alleles are passed to the recipient through a split bottleneck of multiple particles; as a result, the donor and recipient share the same within-host variant, but the consensus allele does not change. In **(F)**, an iSNV arises at site *k* in the donor, and both alleles are passed to the recipient through a split bottleneck of multiple particles; as a result, the donor and recipient share the same within-host variant, and the consensus allele changes.

## RESULTS

### Sub-Consensus Variant Frequencies Exhibit a Power-Law Distribution

To study the distribution of minor allele frequencies, we developed a mathematical model of the viral population within a host (see **Supplement A** and [25–27]). The model consists of two phases: one of *exponential growth* of the inoculum, followed by a *quiescent* phase that lasts from when the viral population reaches a sufficiently large size until the infection ends. We model within-host viral replication during the first phase as a stochastic pure-birth process, consistent with previous methodological work [28], in which each birth event may introduce a *de novo* mutation with some probability *p*. Under the pure-birth model, mutations that emerge early in infection constitute a larger fraction of the ultimate viral population than those that emerge later in infection. Once the within-host viral population size is sufficiently large (see **Supplement B** for a precise mathematical definition), any mutation events that occur negligibly affect the composition of the viral population. Therefore, following the exponential growth of the pure-birth model, we model the quiescent phase via the Jukes-Cantor model [29] of neutral evolution. The combination of these two phases yields a biologically-motivated hierarchical model for within-host variant frequencies in the absence of natural selection. Per **Supplement B**, the probability density function of the frequency *x* of a *de novo* minor variant is approximately proportional to 1/*x*^2^, for any sufficiently small value of *p*.

To assess the validity of our model for *de novo* minor variant frequencies, we visualized all iSNV frequencies observed across a dataset of 134,682 genomes, applying a 3% minor allele frequency threshold [2]. We observed minimal discrepancy between the empirical probability density and the theoretical probability density function, *f*(*x*) ∝ 1/*x*^2^ for *x* > 3% (**Figure 2**). We estimated the mutation rate to be 3.40 × 10^-6^ nt site^-1^ cycle^-1^ by maximum likelihood, comparable to existing estimates of 1–3 × 10^-6^ nt site^-1^ cycle^-1^ [24,30–32] (see **Methods**). Given a substitution rate of 1 × 10^-3^ nt site^-1^ year^-1^ [30–32], our estimate corresponds to a within-host replication rate of 0.81 cycle day^-1^. Per Bar-On et al.’s estimate of a burst size of ∼1,000 particles, we estimate that it takes 2.48–3.72 days for a host to reach a peak viral population of 10^6^–10^9^ virions respectively, in concordance with existing estimates of approximately 3 days [30].

**Figure 2:**
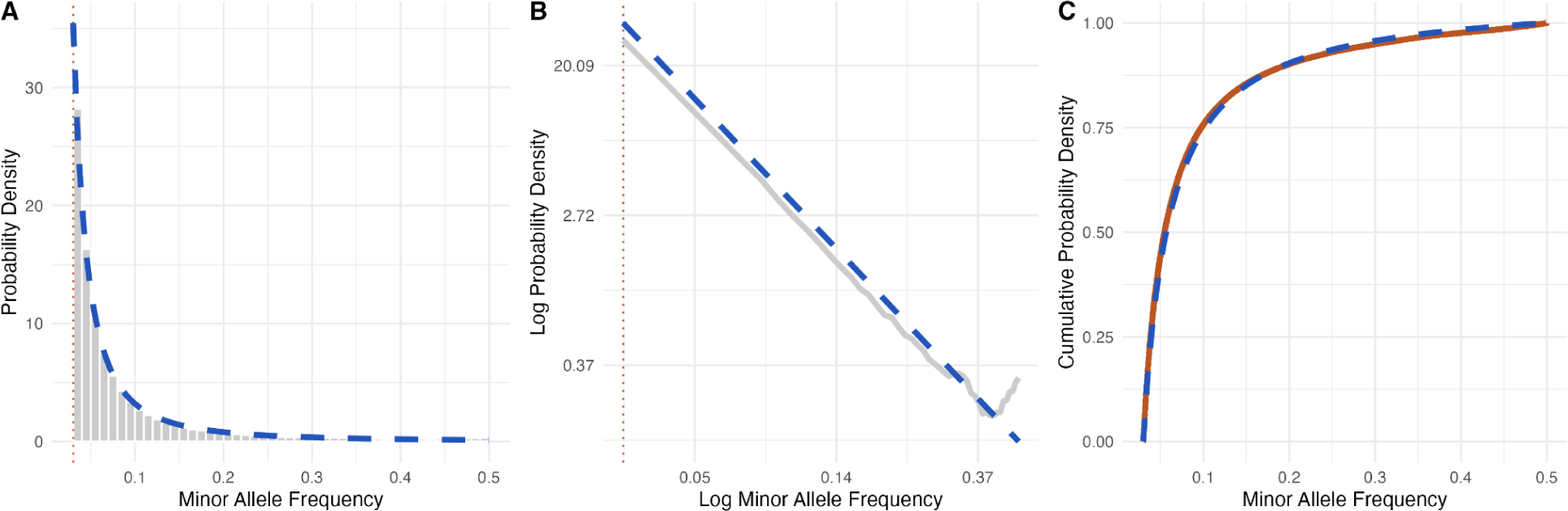
**(A)** Histogram of minor allele frequencies from the dataset of 134,682 genomes (gray bars) and modeled probability density function (blue dashed line); **(B)** Empirical cumulative density function (orange solid line) and modeled cumulative density function (blue dashed line). A minor allele frequency filter of at least 3% was applied for both figures; **(C)** Log-log plot of empirical (gray) and theoretical density functions.

### Within-Host Variants Improve Transmission Inference Accuracy

Extending our model for within-host evolution to account for transmission events gives rise to a model for viral outbreaks as a whole. To quantify the likelihood of any possible transmission bottleneck, we modeled the probability of any individual virion passed from one host to another exhibiting an A, C, G, or T at a given site as a categorical draw, independent and identically distributed across sites on the viral genome and across transmitted particles (see [28] for comparable methods). We combined this genomic likelihood function with a standard epidemiological likelihood function for the susceptible-exposed-infectious-removed (SEIR) stochastic-epidemic process, yielding a Bayesian statistical model of outbreaks as a whole (see **Methods**).

To assess the accuracy of our outbreak reconstruction model relative to existing ones, we first generated a dataset of 100 synthetic outbreaks. We modeled the generation intervals, latent period durations, and infectious period durations as a stochastic-epidemic process, with each time interval independently sampled from a pre-specified probability distribution based on existing studies of SARS-CoV-2 Omicron variant transmission [33,34] (see **Figure 3A** and **Methods**). We simulated transmission networks by generating random trees in which the number of offspring of each case follows a negative binomial distribution with a basic reproductive number (*R*_0_) of 2.5 and overdispersion parameter (*k*) of 0.1 [35], and extracting a random subtree of 10–15 individuals to be our cluster, discarding trees in which no such subcluster existed. We also included an additional 0–5 cases from a different part of the tree in our cluster who each tested positive within 10 days of someone in the subtree, allowing us to test our model’s ability to identify distinct clusters.

**Figure 3:**
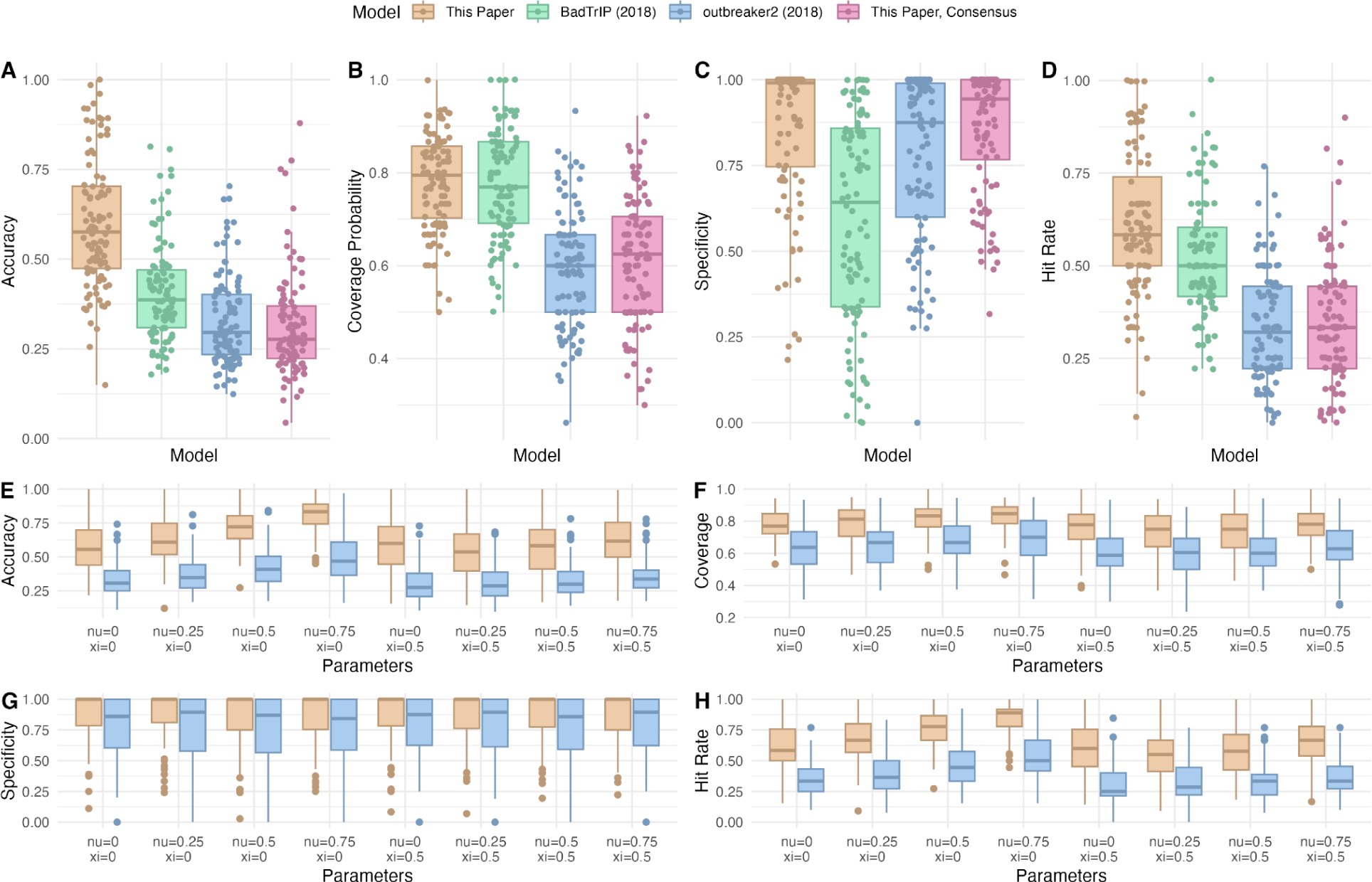
**(A), (B), (C), (D)** Boxplots of accuracy, coverage probability, specificity, and hit rate for our model, outbreaker2, our model using only consensus genomes, and BadTrIP. **(E), (F), (G), (H)** Boxplots of accuracy, coverage probability, specificity, and hit rate for our model and outbreaker2 under eight parameter combinations for contact tracing, where *ν* is the true positive rate and *ξ* is the false positive rate.

We generated synthetic genome sequencing data to reflect the evolution of SARS-CoV-2 as closely as possible. We sampled minor allele fractions at each site for each host per the distribution shown in **Figure 2**, with the probability of a given site exhibiting a SNV (or iSNV) proportional to the number of times we detected the SNV at the given site among the 134,682 Massachusetts genomes. We sampled the virions passed from donor to recipient as independent, identically distributed (i.i.d.) categorical random variables taking on one of four values corresponding to the four nucleotides, per the methods of Leonard et al. [28]. We set the per-site, per-cycle mutation rate to 3.40 × 10^-6^, and assumed a per-site, per-year substitution rate of 1 × 10^-3^ [30–32]. These two parameters are related to each other through the burst size and replication rate, whose values were assigned based on existing literature [24,30] (see **Methods**).

We measured the reliability of our outbreak reconstruction model using four metrics: accuracy, coverage probability, specificity, and hit rate. *Accuracy* is defined as the probability that a random true donor-recipient pair is present in a given posterior sample. *Coverage probability* is defined as the chance that a minimal 90%-credible set^1^ of modeled donors for a given recipient contains the correct donor (or correctly identifies the recipient as unrelated to the cluster).

*Specificity* is defined as the probability that in a given posterior sample, a random unrelated case is correctly identified as being disconnected from the cluster. Finally, *hit rate* is defined as the chance that the most probable ancestor for each case as inferred by our algorithm is indeed the true ancestor.

We compared these metrics of reliability for our method, as well as our method adapted to analyze only consensus genomes, to two leading transmission reconstruction methods: outbreaker2 and BadTrIP. Our method using read-level data outperformed the other three by all four metrics, exhibiting 18.9% higher accuracy than any other tool (see **Table 1** and **Figure 3A–3D**.) **Figure 4** provides an illustrative example of the differential performance of our method and outbreaker2 on a single synthetic outbreak.

**Table 1:** Performance metrics of our model, outbreaker2, BadTrIP, and our model using only consensus genomes as an input.

| *Model* | *Accuracy (SD)* | *Coverage Prob. (SD)* | *Specificity (SD)* | *Hit rate (SD)* |
| --- | --- | --- | --- | --- |
| This paper | 59.9% (18.1%) | 78.0% (10.3%) | 85.9% (21.0%) | 61.0% (19.5%) |
| BadTrIP | 41.0% (13.8%) | 77.8% (11.6%) | 59.8% (30.3%) | 51.9% (16.3%) |
| outbreaker2 | 33.0% (12.6%) | 60.0% (13.1%) | 77.1% (24.5%) | 33.1% (14.9%) |
| This paper,<br>consensus<br>genomes | 31.5% (14.4%) | 61.2% (13.7%) | 85.3% (17.9%) | 34.4% (16.7%) |

**Figure 4:**
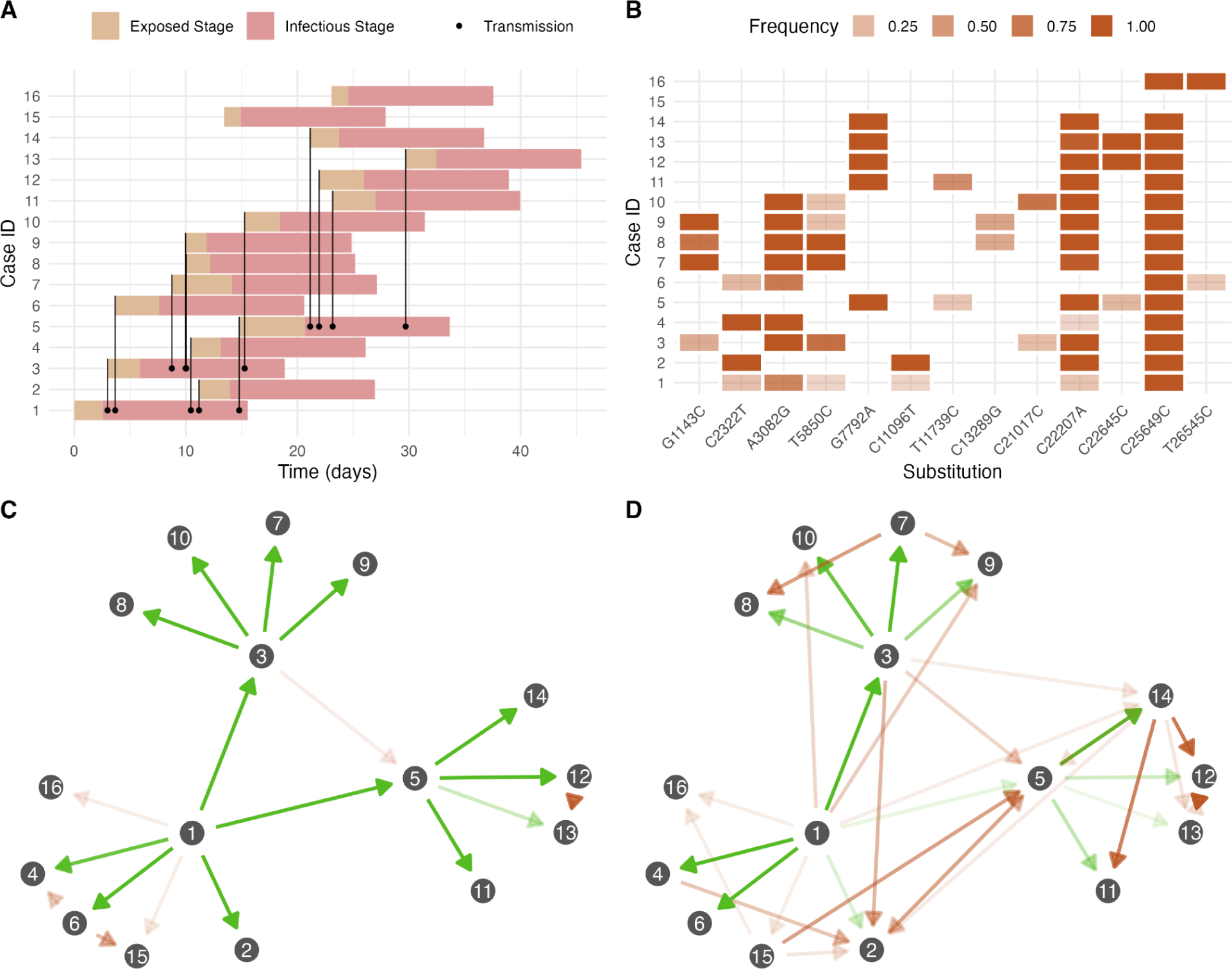
**(A)** An example synthetic outbreak, showing the latent (orange) and infectious (red) stages for each case, as well as transmission events (black lines originating from circles). **(B)** Genetic profile of a synthetic outbreak, showing iSNVs and fixed SNVs for each case (mutations appearing in at most one case not shown). **(C)**, **(D)** Transmission networks for the example synthetic outbreak as inferred by our model and by outbreaker2 (2018), respectively, with the opacity (alpha) of each transmission link corresponding to its posterior probability. A green arrow indicates a correct transmission link, i.e., one that exists in the synthetic outbreak, and an orange arrow indicates an incorrect transmission link. Cases 15 and 16 were not part of the same cluster as cases 1–14.

Given that some outbreak reconstruction models, including our model and outbreaker2, can integrate contact tracing into transmission inference, we further benchmarked performance with this additional form of data. We ran our model and outbreaker2 on the 100 synthetic datasets under eight different contact tracing scenarios: true positive rate *ν* (probability of reporting a contact, given that a transmission occurs) equal to 0%, 25%, 50%, or 75%; and false positive rate *ξ* (probability of reporting a contact, given that a transmission does not occur) equal to 0% or 50%. False-positive contact-traced links were sampled from all pairs of people for which a transmission did not occur. Our model significantly outperformed outbreaker2 in terms of accuracy, coverage probability, and specificity in all scenarios (see **Figure 3E–3H**).

### Animal Study and Contact-Traced Outbreak Validate Transmission Inference Method

Validating our ability to detect transmission events from genomic data alone is hindered by the lack of controlled experiments or well sampled outbreaks with known transmission routes.

Nonetheless, we identified one controlled study of BRSV in Swedish cows [18] and one nosocomial outbreak in South Africa [36,37] as strong candidates for methodological validation on real data. We chose to reconstruct the BRSV outbreak because it was a controlled study with known infection dynamics, and because the pathogen exhibited comparable substitution rate to SARS-CoV-2, estimated at 1.6 × 10^-3^ nt site^-1^ yr^-1^ for synonymous mutations [38]. In the BRSV outbreak, transmissions occurred either through co-housing or by obtaining bronchoalveolar lavages from infectious animals and using them to infect naïve animals, with each successive transmission event occurring seven days after the previous one. All cows were housed in separate pens (with the exception of the aerosol transmission step), to avoid unintended transmission pathways [18]. The transmission network is shown in **Figure 5A**. We reconstructed the BRSV outbreak using our tool, BadTrIP, outbreaker2, and our tool with only consensus genomes (see **Figure 5B–5E**). The four approaches yielded accuracies of 59.3%, 45.5%, 38.9%, and 44.6%, respectively (see **Supplementary Table S1**).

**Figure 5:**
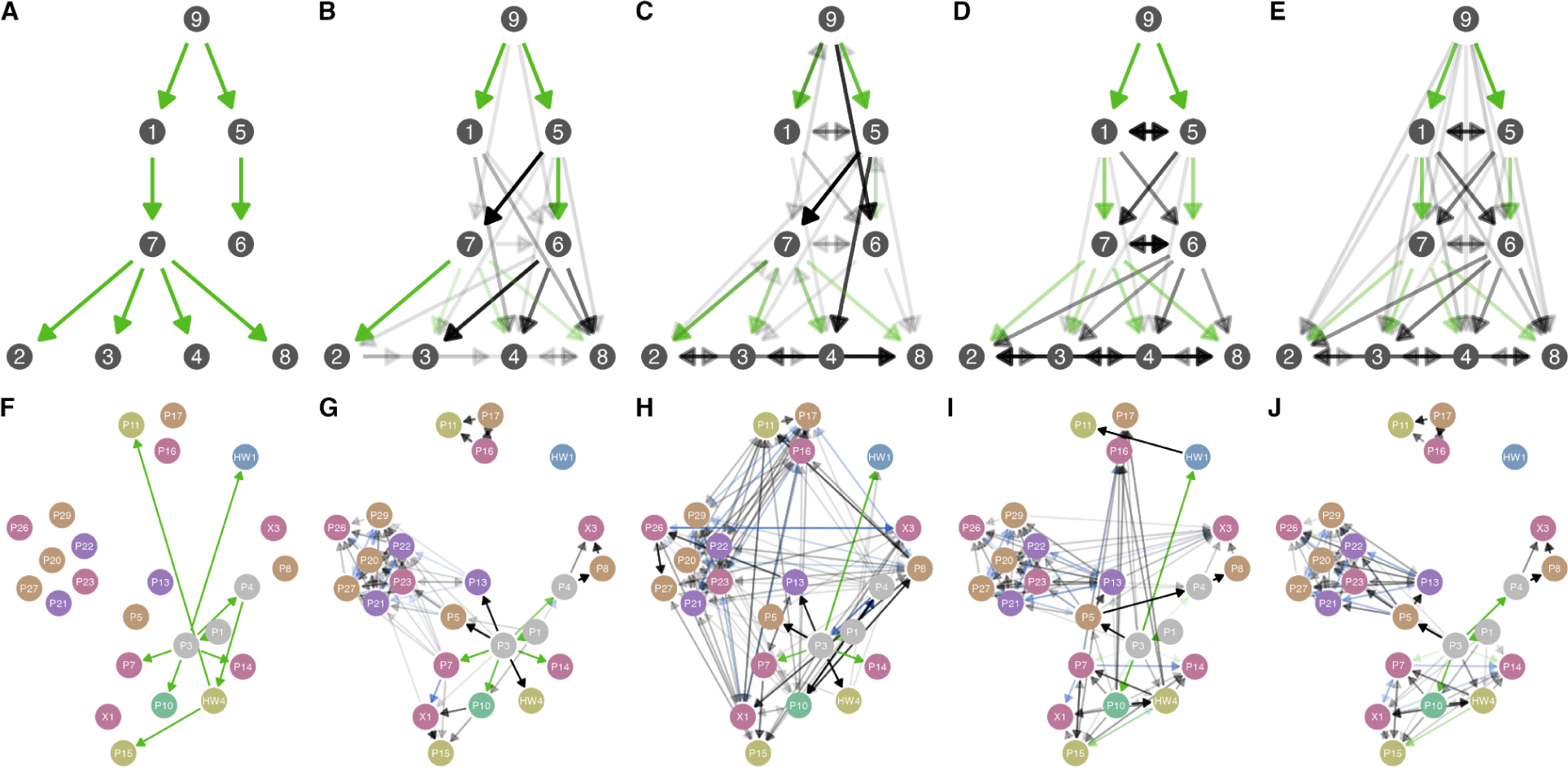
**(A)** Transmission network for the BRSV outbreak. **(B), (C), (D), (E)** Inferred transmission networks for the BRSV outbreak by our tool, BadTrIP, outbreaker2, and our tool with only consensus genomes, with a green arrow representing a correctly-inferred transmission link and a gray arrow representing an incorrectly-inferred transmission link. The opacity of each arrow corresponds to its posterior probability. **(F)** Epidemiologically-confirmed transmissions in the nosocomial SARS-CoV-2 outbreak in South Africa. **(G), (H), (I), (J)** Inferred transmission networks for the SARS-CoV-2 outbreak by our tool, BadTrIP, outbreaker2, and our tool with only consensus genomes, with a green arrow representing an epidemiologically-confirmed transmission link, a blue arrow representing a plausible transmission link based on the location histories of each patient, and a gray arrow representing a transmission link with neither form of supporting evidence. The opacity of each arrow corresponds to its posterior probability. Nodes are color-coded by facility: orange represents the medical intensive care unit, red represents a medical ward, green represents a nursing home, blue represents the cardiac intensive care unit, purple represents the surgical intensive care unit, yellow represents the neurology ward, and gray represents patients who traveled between different wards.

We then applied the same four methods to reconstruct a SARS-CoV-2 outbreak at a hospital in South Africa between March and April of 2020 [36,37]. We selected this outbreak because among the 39 infected persons, 9 transmission links between patients with sequenced samples were deemed highly probable by epidemiological investigation (see **Supplementary Table S3**), and the COVID-19 prevalence was comparatively low at the time in South Africa (< 0.02% nationally [39]) as to reduce the probability of undetected external introductions. In addition to epidemiologically-confirmed links, patient movement histories throughout the hospital were documented, suggesting certain transmissions to be plausible based on patient co-location. Our tool, BadTrIP, outbreaker2, and our tool with only consensus genomes, inferred the epidemiologically-confirmed transmission links with accuracies of 49.5%, 35.1%, 36.8%, and 36.5%, respectively (see **Figure 5F–J** and **Supplementary Table S2**).

### Outbreak Reconstruction Identifies Genetic Signatures of Transmission

Outbreak reconstruction with read-level sequencing data allows us to assess the prevalence and impact of informative within-host diversity. To do so, we extracted putative outbreak clusters from the 134,682 genomes by creating a network graph, in which a node represents one patient, and an edge connects two patients if and only if their sequenced samples exhibited a SNV distance of at most 2 and were collected less than 10 days apart. We then defined a cluster as a connected component of this graph with at least two nodes. This filtering process identified 5,692 putative clusters with *n* ≥ 2 individuals (38,598 individuals total), with median size of 3 individuals per cluster (interquartile range: 2–5). The mean and standard deviation were 6.78 and 14.3, respectively, indicating substantial overdispersion as is typical of outbreak sizes. We ran our reconstruction algorithm on each cluster, and tabulated all putative transmission links and their associated posterior probabilities across all clusters. We found that in a random posterior sample, a randomly-selected transmission link has a 12.3% probability (95% CI via bootstrap: 11.6%–13.0%) of exhibiting at least one genetic relationship visible only in read-level data. These relationships, which we refer as the *three iSNV signatures*, are: a minor allele in the donor becomes fixed in the recipient (minor-to-fixed), a fixed allele in the donor becomes the minor allele in the recipient (fixed-to-minor), or the major and minor allele at a polymorphic site in the donor are both present in the recipient (split bottleneck). The other 81.1% of links exhibited no informative within-host diversity.

While the three iSNV signatures appear in the minority of transmission links, they are disproportionately useful for resolving transmission networks. To see this more concretely, we first considered putative transmission links in which the SNV distance between consensus genomes equals 0 (62.6% of putative links). The only possible iSNV signature such a transmission link may exhibit is a split bottleneck. Among transmission links with 0 SNV distance, the posterior probability equalled 41.7% on average (95% CI via bootstrap: 40.5%–42.9%) in the presence of a split bottleneck, and 18.4% otherwise (95% CI via bootstrap: 18.3%–18.6)^2^. Transmission links with a nonzero SNV distance (37.4% of putative links), on the other hand, may exhibit any of the three iSNV signatures. Although the posterior probabilities associated with these links did not differ significantly based on the presence of an iSNV signature, we found that 24.4% of them exhibit an iSNV signature (95% CI via bootstrap: 23.1%–25.5%). In other words, 24.4% of the time, a transmission link with nonzero SNV distance could be better inferred with read-level data as compared to consensus genomes, highlighting one central way that our transmission reconstruction tool improves upon existing methods.

## DISCUSSION

Here, we establish a novel method for modeling genetic diversity in outbreaks, providing a systematic approach for inferring transmission links from read-level sequencing data. By modeling the distribution of iSNV frequencies, we quantified the likelihood of any possible viral outbreak transmission network based on the genomic and epidemiological data collected. Synthetic outbreak simulations demonstrated that using read-level data in outbreak reconstruction significantly improved accuracy, coverage probability, specificity, and hit rate compared to using consensus genomes alone, as our model was able to detect minor allele transmissions and split bottlenecks. Moreover, as we show, while BadTrIP offers some improvement in accuracy over consensus-based methods, accuracy can be improved much further with our approach to modeling minor variants. Our methods, applied to 5,692 outbreak clusters, demonstrate the critical role that minor variants play in resolving transmission links.

Our findings have implications across virology, epidemiology, and genetics. Our model for minor allele frequencies alone provides a reliable means of estimating the per-site, per-cycle mutation rate without needing to calculate it from the substitution rate per-unit-time, which requires knowledge of the viral population growth rate and the number of virions produced per cycle. Moreover, outbreak reconstruction has numerous applications to public health, such as estimating the reproductive number and overdispersion parameter, estimating attack rate by vaccine status, investigating the origin of a cluster, and identifying likely viral transmission pathways in nosocomial settings. With our tool, we provide a more accurate and interpretable method for conducting these analyses.

However, our outbreak reconstruction model faces a number of limitations in its current form. First, the relatively slow substitution rate of viruses like SARS-CoV-2 limits the resolution of any genomics-based outbreak reconstruction method because, in any given outbreak, we expect to see a number of cases with identical consensus genomes and no informative within-host diversity. In particular, lack of informative within-host diversity limits our ability to differentiate between multiple introductions of identical lineages versus within-facility transmission, negatively impacting the accuracy of our tool (mean = 59.9%, SD = 18.1% for synthetic datasets). While we outperformed existing tools, we emphasize that we would not expect any tool to perfectly reconstruct transmission networks for any virus with a relatively low mutation rate such as SARS-CoV-2. Second, our tool in its current state does not account for unsampled intermediates: for instance, our tool might reconstruct a transmission chain A → B → C where A and C are sequenced, but B is not, as A → C. This limitation could conceivably be addressed by modifying the MCMC algorithm to add and remove unsequenced cases. Finally, obtaining reliable and reproducible iSNV calls via deep sequencing remains a challenge, and the detection of within-host variants may be biased by the method of sample collection (e.g., nasopharyngeal swab, oral swab) and by the time of collection relative to symptom onset. The presence or absence of an informative iSNV in a given sample may significantly influence the output of our outbreak reconstruction tool.

Our model also makes a number of simplifying assumptions about the biological processes underlying viral infection and transmission, which is necessary to reduce computational complexity. In line with other outbreak reconstruction models, we assume that mutations are neutral. This simplification fails to account for cases in which an iSNV rises to a detectable frequency due to a selective advantage, such as immune escape, or is lost due to either genetic drift or a selective disadvantage. However, we do our best to minimize the effect of this simplification on our reconstruction of SARS-CoV-2 outbreaks by quantifying the frequency of specific iSNVs across our 134,682 genomes and masking recurrent iSNV calls. In addition, our model assumes that sites evolve independently, ignoring epistatic effects. It also assumes that the intrahost viral population is well-mixed, without spatial heterogeneity in genotype. Finally, in this paper, we tailor our approach to single-stranded RNA viruses. Extensions of our model would be required to account for more complex pathogens, such as those with extensive recombination or horizontal gene transfer.

Despite these challenges, our work underscores the usefulness of read-level viral genome data and high-quality iSNV calls in transmission inference. At present, within-host variant calls below 3% frequency are typically considered unreliable [2], though iSNVs in that range may still be transmitted and provide critical information about putative donor-recipient pairs. Moreover, our work stresses the importance of dense sampling of outbreaks, as missing genomes may lead our model to infer additional index cases or transmissions with large SNV distances erroneously. Given dense sampling and high-resolution sequencing, we expect our method to reconstruct outbreaks with high accuracy.

## METHODS

### Ethical Approvals

The research project (Protocol #1603078) was reviewed and approved by the Massachusetts Department of Public Health (MADPH) Institutional Review Board and covered by a reliance agreement at the Broad Institute. An additional non-human subjects research and an exempt determination (EX-7080) were made by the Harvard Longwood Campus Institutional Review Board and the Broad Institute Office of Research Subject Protections, respectively, for the analysis of de-identified aggregate and publicly available data.

### Analysis of SARS-CoV-2 Sequencing Data

All sequencing data was downloaded from the NCBI SRA under BioProject PRJNA715749. We used LoFreq version 2.1.5 to call intrahost single nucleotide variants (iSNVs). We then filtered out calls with < 100 read depth, < 10 minor allele read count, *p*-value of Fisher’s exact test for strand bias < 0.05. In addition, as a means of masking highly-variable sites, we compiled a table from a larger dataset of 172,519 SARS-CoV-2 genomes sequenced at the Broad Institute listing the number of times an iSNV rising above 3% frequency appeared at each site. From this dataset, we calculated the probability of a randomly chosen iSNV in a randomly chosen host occurring at each site on the viral genome. Based on these probabilities, we masked sites whose probability of being chosen more than once among 1,000 trials exceeded 5%. This method was designed to model the probability of the same iSNV arising *de novo* more than once in an outbreak of 1,000 people.

### Modeling Minor Allele Frequencies

See <u>Supplementary Text</u>, Section A. In brief, we propose a stochastic pure-growth process to model within-host viral replication, where each birth event may introduce a given substitution with some probability. If the substitution probability is small, then we can approximate the probability density function (PDF) of the proportion of mutated particles by the function 1/*x*^2^.

### Outbreak Reconstruction Model

See <u>Supplementary Text</u>, Section B. In brief, we develop a statistical model for the proportion of the within-host viral population to exhibit a given nucleotide at a given site, for any host at any point in time. This model combines the result from Supplementary Text A with the standard Jukes-Cantor model of neutral evolution. We then combine this genetic model with an epidemiological model, in which each host may be susceptible, exposed, infectious, or recovered at any given time. Together, these two components provide an overall likelihood function for viral outbreaks with read-level sequencing data.

### MCMC Implementation

See <u>Supplementary Text</u>, Section C. In brief, we propose several moves to search the space of possible transmission networks efficiently, as well as standard moves to update underlying parameter values.

### Synthetic Data Generation

We simulated outbreaks by first generating stochastic transmission networks over the course of four generations. We started with a single index case, and assumed a negative binomial offspring distribution with mean *R*_0_ = 2.5 and overdispersion parameter *k* = 0.1. We then overlaid a stochastic-epidemic process identical to that in our outbreak reconstruction model (see <u>Supplementary Text</u>, Section B.4). Next, we selected a fully-connected subtree with 10–15 nodes uniformly at random, discarding iterations for which no such subtree existed. We also included 0–5 nodes not in our subtree who tested positive at most 10 days before or after the first or last case in the subtree, respectively, to simulate unrelated introductions in an outbreak. Finally, we overlaid the same evolution model as in our outbreak reconstruction algorithm (see <u>Supplementary Text</u>, Section B.4). We assumed that the sequencing coverage for every case was 100%, with read depths ranging from 100 to 20,000, uniformly at random. All parameter values are equal to those listed in <u>Supplementary Text</u>, Section C.1.

## Data Availability

The outbreak reconstruction tool is available as an R package on GitHub, at github.com/broadinstitute/reconstructR. Code and data for all analyses and figures in this paper are available at github.com/broadinstitute/transmission-inference. All sequencing data used in this study are publicly available through the NCBI SRA under BioProject PRJNA715749.

https://github.com/broadinstitute/transmission-inference

## SUPPLEMENTARY TABLES

**Supplementary Table S1:** Accuracy, coverage probability, and hit rate of the four methods applied to the BRSV outbreak.

| *Model* | *Accuracy* | *Coverage Prob.* | *Hit Rate* |
| --- | --- | --- | --- |
| This paper | 59.3% | 8 of 9 | 5 of 9 |
| BadTrIP | 45.5% | 7 of 9 | 4 of 9 |
| outbreaker2 | 38.9% | 9 of 9 | 4 of 9 |
| This paper, consensus genomes | 44.6% | 9 of 9 | 4 of 9 |

**Supplementary Table S2:** Accuracy, coverage probability, and hit rate of the four methods applied to the South Africa SARS-CoV-2 outbreak.

| *Model* | *Accuracy* | *Coverage Prob.* | *Hit Rate* |
| --- | --- | --- | --- |
| This paper | 49.5% | 5 of 9 | 5 of 9 |
| BadTrIP | 35.1% | 4 of 9 | 3 of 9 |
| outbreaker2 | 36.8% | 4 of 9 | 3 of 9 |
| This paper, consensus genomes | 36.5% | 5 of 9 | 4 of 9 |

**Supplementary Table S3:** Epidemiological evidence supporting the putative transmission links identified in the South Africa SARS-CoV-2 nosocomial study.

| *From* | *To* | *Supporting Evidence from [36]* |
| --- | --- | --- |
| P1 | P3 | “On review of the timeline, it was noticed that the first outpatient case (P1) and the person who would become the first inpatient case (P3) were both in the Emergency Department on 9 March. A more detailed investigation of events of that day uncovered that they were in the ED at overlapping times, were in close proximity to one another, and were attended to by the same medical officer.” |
| P3 | HW1 | “On review of [HW1’s] shift patterns and patient allocation, it was noted that she was directly responsible for the care of P3 in CICU on the night shift of 12 Mar–13 March. Based on the understanding that people can be infectious 1–3 days prior to symptom onset, this link provides strong circumstantial evidence that P3 was infectious at this time and transmitted |
|  |  | to the CICU nurse.” |
| P3 | P4, P7, P14 | “On the assumption that P3 was infectious throughout her time on MW1 (13-16 Mar), there were five other cases who may have been exposed during that time period. This includes a 46-year-old female (P4) who was in the bed directly opposite (in room 12), and four male patients (P6, P7, P14, X4) who were co-located in a room down the corridor (room 15).”<br>[Note: P6 and X4 were not sequenced.] |
| P4 | HW4 | “A professional nurse on this ward (HW4) became symptomatic with fever and flu-like symptoms at the end of her shift on 23 March and subsequently tested positive on 29 March. This nurse worked directly with P4 on 23 March, when P4 was coughing, and performed tasks including nebulisation.” |
| HW4 | P11, P15 | “[HW4] was signed off work until 27 March and when she returned to work on 27 March she was still symptomatic with cough, fever and sore throat, but she continued to work day shifts on 27 & 28 March. On those days she worked directly with P11 and P15.” |
| P3 | P10 | “On 16 April, patient P3 was discharged from St. Augustine’s Hospital to the Bill Buchanan Association for the Aged in Morningside, a nursing home with 210 residents. She was there until her readmission to St. Augustine’s on 22 March. By 31 March, we understand that four other residents at the home were diagnosed with COVID-19, including three women who had shared the sick bay with P3 and one woman who stayed in a separate unit and only visited P3 (as they were friends from their residential home). One of the cases from the sick bay was admitted to St. Augustine’s on 31 March (patient P10).” |

## CODE AND DATA AVAILABILITY

The outbreak reconstruction tool is available as an R package on GitHub, at <u>github.com/broadinstitute/reconstructR</u>. Code and data for all analyses and figures in this paper are available at <u>github.com/broadinstitute/transmission-inference</u>. All sequencing data used in this study are publicly available through the NCBI SRA under BioProject PRJNA715749.

## ACKNOWLEDGEMENTS

We graciously acknowledge D. Grant, J.D. Sandi, M. Momoh, and the lab at Kenema Government Hospital in Sierra Leone for being early adopters of the methods proposed in this paper; the entire team at Fathom Information Design for their contributions to the interactive visualization of this work; W.P. Hanage, C. Worby, A. Colubri, and K. Koelle for their early feedback on our methods; and I.A. Bhatt for his exploratory research in iSNV phasing, which we hope to incorporate into future iterations of our model.

## AUTHOR CONTRIBUTIONS

Conceptualization, I.O.A.S., P.V., and P.C.S.; data curation, I.O.A.S., G.K.M., and T.B.; formal analysis, I.O.A.S., B.A.P., G.K.M., T.B., L.A.K., and P.V.; funding acquisition, P.C.S.; investigation, I.O.A.S., B.A.P., G.K.M., T.B., and P.V.; methodology, I.O.A.S., B.A.P., G.K.M., T.B., M.D.M., and P.V.; project administration, I.O.A.S., P.V., and P.C.S.; resources, B.A.P., G.K.M., T.B., L.A.K., and P.V.; software, I.O.A.S. and P.V.; supervision, S.F.S., D.J.P., B.L.M., P.V., and P.C.S.; visualization, I.O.A.S., M.S., K.Y., O.G., and B.F.; writing – original draft, I.O.A.S.; writing – review & editing, I.O.A.S., B.A.P., G.K.M., T.B., L.A.K., D.J.P., B.L.M., M.D.M., P.V., and P.C.S.

1 By “minimal 90%-credible set” we mean the smallest set of possible donors for a given recipient whose combined probability of being the true donor exceeds 90%.

2 Averages and confidence intervals computed after removing transmission links with <1% posterior probability, to remove large volumes of highly improbable transmissions.

