## Supplementary Text for "Inferring Viral Transmission Pathways from Within-Host Variation"

#### A Modeling Minor Allele Frequencies

##### A.1 Stochastic Pure-Birth Process with Mutations

We model viral RNA replication within the host as a *stochastic pure-birth process*. This term refers to a model of a population—here, a population of virions—in which birth events, i.e. events in which existing virions replicate to form new ones, occur randomly over time. Each birth event may result in one or more offspring, and the rate at which birth events occur may depend on the current population size. Moreover, the term *pure-birth* means that the population size only ever increases. Such a model is reasonable for the early days of an infection, when the viral population within a host grows much faster than it declines.

In our model, there are $a$ virions within a host at time 0, where $a$ is a positive integer. We then model the times at which an individual virion births new virions as following a *Poisson process* with rate $\lambda$. This means that if a given virion $v$ is birthed at time $T_0$, and virion $v$ gives birth at times $T_1, T_2, T_3, \dots$, then the intervals between the births of the offspring of virion $v$, i.e. $T_1 - T_0, T_2 - T_1, T_3 - T_2, \dots$ are independent and identically distributed $\text{Expo}(\lambda)$ random variables. We model birth events as instantaneous, and assume that the number of offspring of a given birth event always equals a positive integer $w$.

Finally, to capture mutations, we add one additional layer of complexity to our model. Suppose that each site on the viral genome can be white or black, which we take to represent whether or not a virion exhibits a mutation at the given site. Suppose that the colors of the intial $a$ virions at time 0 are known. Then for each birth event, we model the color of the $w$ offspring virions as being the opposite color as the parent with probability $p$, and the same color as the parent with probability $1 - p$. All offspring for a given birth event always exhibit the same color. Note that under this model, we allow for only one possible mutation at each site within each host; as such, our model does not account for multiallelic sites.

##### A.2 Biological Applicability

The above stochastic pure-birth process aptly models the process of a virus replicating within a host, under certain assumptions. The number $a$ describes the number of viral particles that inoculate the host, also referred to as the transmission bottleneck size. The number $w$ is the burst size, i.e. the number of ofspring per birth event. In reality, this number may vary between cell infection events. In our model, however, we assume every cell infection produces the same number of offspring virions, but leave $w$ as an unknown parameter to be inferred by the model. Moreover, our model assumes that birth events are instantaneous as a simplifying assumption. In reality, the total duration of the intracellular replication process is estimated at about 7–8 hours for a SARS-like virus (Sender et al. 2020).

Finally, we assume that if a mutation occurs at a site, it affects all offspring particles at that site. When an infectious virion enters a cell, a "template strand" of RNA is synthesized, equal to the reverse complement of the virion's genetic material. Then, numerous new copies of RNA are produced from the template, identical to the genetic material of the original virion, except possibly with some mutations. This entire process of producing a template from which to create new genetic material may only occur once, or may recur multiple times. While still an open area of biological research, there is limited evidence suggesting that for some RNA viruses such as influenza, the replication process typically involves producing one or few templates from which all offspring viral RNA is produced, as opposed to numerous templates that each produce a fraction of the overall viral RNA output from the cell (Sanjuán et al. 2010). In our model, we assume only one template RNA is created, from which all offspring are produced.

Under this assumption, the step at which a mutation occurs—that is, whether during the creation or the copying of the template—is very important. If a mutation occurs when copying the virion that enters a cell to produce a template, then approximately all offspring virions exhibit that mutation. If a mutation occurs when copying the template RNA to produce offspring virions, then we would expect that mutation to appear in only one offspring. In our model, we assume the latter scenario negligibly impacts the overall fraction of virions exhibiting a given mutation, since a typical burst size ($w$) tends to be on the order of $10^1$ to $10^3$ (Sender et al. 2020), and a mutation that occurs in the template-copying step would affect only a single virion. To test the validity of this assumption, we simulated the genetic composition of virions within a host using two models, one in which mutations may occur when copying the template RNA to produce offspring virions, and the other in which they may not. Assuming an final viral population size of $10^6$, an initial viral population size of 1, a burst size of 500, and a mutation rate of $3 \times 10^{-6}$ substitutions per site per cycle, we found the two models to exhibit very similar PMFs (see Figure 1).

#### A.3 Distribution of Mutated Particles

If we assume mutation events are rare, then it is possible to derive an approximate distribution for the fraction of virions exhibiting a given mutation after $k$ birth events. Since birth events occur randomly in continuous time, we may assign them an order. That is, we may refer to the 1st, 2nd, 3rd, ... birth events to occur over time unambiguously, despite the fact that for a growing population, these events will happen closer and closer together. One very convenient property of our model is the following: Suppose we freeze the pure birth process at a time in which there are $m$ viral particles, and label the particles $1, 2, \ldots, m$. Then the *next* particle to give birth is uniformly distributed over the set of integers $1, 2, \ldots, m$, i.e. all possible parents are equally likely. Intuitively, this follows from the fact that the Exponential distribution is memoryless, i.e. at any point in time $t$, the interval between $t$ and the next time a given virion $v$ gives birth still follows an Expo($\lambda$) distribution (for a rigorous proof, see Specht and Mitzenmacher (2023), Section 5).

The property that at any given time, the next particle to give birth is uniformly distributed, is important because it provides us with a mathematical tool to study the distribution of black and white (non-mutated and mutated) particles after some number $k$ of birth events. This mathematical tool is called a *generalized Pólya Urn model with random replacement*, the details of which may be found in Specht and Mitzenmacher (2023). One useful property of the generalized Pólya Urn model

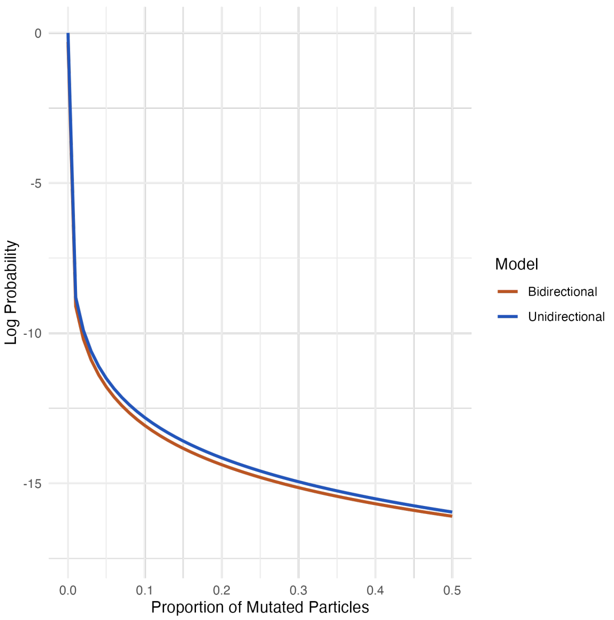


Figure 1: PMF of the minor allele fraction where mutations may occur both when creating the template and copying the template to produce offspring virions (orange, “bidirectional”), versus only when creating the template (blue, “unidirectional”). Note: probabilities are plotted on a log scale.

is as follows: Suppose at some time $t_0$, $u + v$ birth events have occurred, where $u$ of the birth events produce white particles, and $v$ of the birth events produce black particles. Suppose at a later time $t_1$, the viral population has increased by some amount $c$. Then for large $c$, the fraction of the viral population that consists of white particles approximately follows a $\text{Beta}(u, v)$ distribution. Specifically, the $\text{Beta}(u, v)$ distribution is the limiting distribution of this scenario as $c \to \infty$ and $p \to 0$. This use of the Beta distribution has previously been applied to numerous studies of within-host variation—notably Leonard et al. (2017) in their estimation of transmission bottleneck sizes.

So, let us use this Beta distribution property to generate a mathematical model for the fraction of the viral population to exhibit a given mutation, after $k$ birth events have occurred. Per our white-and-black particle notation, this is equivalent to modeling the fraction of virions that are colored white after $k$ birth events. Let $X$ be a random variable denoting this fraction, and let $g_{k,u,v} : [0, 1] \to \mathbb{R}$ denote its probability density function, where $u$ and $v$ are the numbers of black and white virions at time 0, respectively. First, we note that $g$ will depend—very significantly, in fact—on $u$ and $v$. To address this, we will divide our model into two cases: (1) some of the initial $a$ particles are black, and some are white ($u > 0$ and $v > 0$); and (2) all of the $a$ particles are black ($u = 0$) or all of the $a$ particles are white ($v = 0$).

In the first case, where $u > 0$ and $v > 0$, the work of Leonard et al. (2017) already provides us with a distribution for $X$. The generalized Pólya Urn property tells us that, approximately, the distribution of $X$ is $\text{Beta}(u, v)$. Let $f_{\text{Beta}}(x; u, v)$ denote the probability density function (PDF) of the Beta distribution with parameters $u$ and $v$. Then we write:

$$g_{k,u,v}(x) = f_{\text{Beta}}(x; u, v) \quad \text{when } u > 0 \text{ and } v > 0.$$

In the second case, however, the above equation no longer applies—the reason being that if $u = 0$, then $X$ is deterministically 0, and if $v = 0$, then $X$ is deterministically 1 under the Leonard et al. model. In reality, even if none of the inoculating viral particles for a host exhibit a given mutation, said mutation may still arise *de novo*. To account for this, we invoke our assumption that $p$, the probability of a mutation occurring in a given birth event, is small relative to $1/k$. (In supplement B, we discuss what constitutes a reasonable range of values for $k$, and why the assumption $p << 1/k$ holds for such values.) Under this assumption, with high probability, the number of birth events with a mutation is 0 or 1. If 0 mutations occur among the first $k$ birth events, then $X$ is deterministically 0 in the case $u = 0, v = a$ and deterministically 1 in the case that $u = a, v = 0$. The probability of 0 mutations occurring among the first $k$ steps is $(1 - p)^k$.

The final remaining case, in which 1 mutation occurs among the first $k$ birth events, requires slightly more machinery. First, we approximate the probability of 1 mutation occurring as $1 - (1 - p)^k$, i.e. the probability that more than 0 mutations occur. We then condition on the birth event $M$ at which the mutation occurs. By the property of the generalized Pólya Urn model, if we started with all black particles at time 0 ($u = 0, v > 0$) and the $M$th birth event produces white particles, then the distribution of $X$ is approximately Beta$(1, M)$, conditional on $M$. Symmetrically, if we started with all black particles at time 0 ($u = 0, v > 0$) and the $M$th birth event produces white particles, then the distribution of $X$ is approximately Beta$(M, 1)$, conditional on $M$. Using the fact that $M$ is uniformly distributed over birth events $1, 2, \ldots, k$, we perform the following calculation to marginalize $X$:

$$g_{k,u,v}(x) = \begin{cases} \frac{1-(1-p)^k}{k} \sum_{m=1}^{k} f_{\text{Beta}}(1, m), & u = 0, v > 0 \\ \frac{1-(1-p)^k}{k} \sum_{m=1}^{k} f_{\text{Beta}}(m, 1), & u > 0, v = 0 \end{cases}$$

$$= \begin{cases} \frac{(1-(1-p)^k)(1-(1-x)^k(1+kx))}{kx^2}, & u = 0, v > 0 \\ \frac{(1-(1-p)^k)((k(x-1)-1)x^k+1)}{k(1-x)^2}, & u > 0, v = 0 \end{cases}.$$

In conclusion, the function $g_{k,u,v}$ is defined as follows:

$$g_{k,u,v}(x) = \begin{cases} f_{\text{Beta}}(x; u, v), & u > 0, v > 0 \\ (1-p)^k, & u = 0, v > 0, x = 0 \\ (1-p)^k, & u > 0, v = 0, x = 1 \\ \frac{(1-(1-p)^k)(1-(1-x)^k(1+kx))}{kx^2}, & u = 0, v > 0, 0 < x \leq 1 \\ \frac{(1-(1-p)^k)((k(x-1)-1)x^k+1)}{k(1-x)^2}, & u > 0, v = 0, 0 \leq x < 1 \end{cases}$$

#### A.4 Applying a Minor Allele Frequency Threshold

The function $g_{k,u,v}$ describes the approximate distribution of the fraction of virions exhibiting a given mutation after $k$ birth events. This fraction, in theory, may take on any value ranging from 0 to 1. However, with current genome sequencing technology, detection of minor variants with frequency below a certain threshold (usually 3%) is considered unreliable. In our paper, we report all iSNV calls below 3% as error. Moreover, in our analysis of *de novo* iSNVs across our dataset of 134,682 SARS-CoV-2 genomes, we report minor allele frequencies as the minimum of $f$ and $1 - f$, where $f$ is the frequency detected in the raw deep-sequencing data. We report minor allele frequencies this way because we assume that if an iSNV is indeed *de novo* and its frequency as detected in deep sequencing data is above 50%, then the iSNV was more likely a reversion than a mutation which affects over half of the within-host viral population. Hence, in our *de novo* iSNV analysis, the maximum reported iSNV frequency is 50%.

We account for these practical restrictions on the range of possible reported minor allele frequencies by modifying the probability distribution of $X$. Let $X^*$ be a random variable whose distribution is equal to that of $X$, but with support restricted to $[L, U]$ for real numbers $0 < L < U < 1$. To find the distribution of $X^*$, we first define the following two functions, as a means of simplifying notation:

$$G_k(x) := \int_0^x \frac{1-(1-t)^k(1+kt)}{kt^2}\,dt = \frac{(1-x)^{k+1}+kx+x-1}{kx}$$
$$G'_k(x) := \int_0^x \frac{(k(t-1)-1)t^k+1}{k(1-t)^2}\,dt = \frac{x\left(1-x^k\right)}{k(1-x)}.$$

In addition, let $F_{\text{Beta}}(x; u, v)$ denote the CDF of the Beta distribution with parameters $u$ and $v$ evaluated at $x$. Now that we've defined these functions, let $h_{u,v,k}$ denote the PDF of $X^*$. Then by normalization, $h_{u,v,k}$ is given by:

$$h_{u,v,k}(x) = \begin{cases} \frac{f_{\text{Beta}}(x;u,v)}{F_{\text{Beta}}(U)-F_{\text{Beta}}(L)}, & u > 0, v > 0 \\ \frac{1-(1-x)^k(1+kx)}{(G_k(U)-G_k(L))kx^2}, & u = 0, v > 0 \\ \frac{(1-(1-p)^k)((k(x-1)-1)x^k+1)}{(G'_k(U)-G'_k(L))kx^2k(1-x)^2}, & u > 0, v = 0 \end{cases}$$

for $L \leq x \leq U$. For large $k$ and reasonable values of $L$ and $U$, e.g. $0.001 < L < U < 0.999$, this function is well approximated by:

$$h_{u,v,k}(x) \approx \begin{cases} \frac{f_{\text{Beta}}(x;u,v)}{F_{\text{Beta}}(U)-F_{\text{Beta}}(L)}, & u > 0, v > 0 \\ \frac{LU}{(U-L)x^2}, & u = 0, v > 0 \\ \frac{(1-L)(1-U)}{(U-L)(1-x)^2}, & u > 0, v = 0 \end{cases},$$

with equality as a limit when $k \to \infty$. In our analysis of *de novo* iSNVs across 134,682 SARS-CoV-2 genomes, we assume $u = 0, v = 1, L = 0.03$, and $U = 0.5$, in which case the density of $X^*$ is proportional to the function $1/x^2$ on the domain $[0.03, 0.5]$.

### B Outbreak Reconstruction Model

#### B.1 Preliminaries

Here, we describe the model for viral outbreaks with deep-sequencing data in detail. We first provide an overview of how the within-host viral population may change over time, which will motivate our construction of the model. We then define our unknown parameters and known data. Finally, we define our priors and likelihood function.

As in a typical epidemiological model, we assume each host at any given time may be either susceptible, exposed (i.e. contracted the virus but not yet capable of spreading it), infectious, or recovered (SEIR model). In addition to these epidemiological compartments, we also define two "genetic phases" for each host: the *exponential growth phase*, which starts when the host first contracts the virus and ends when the host reaches a sufficiently large viral population (we will explain "sufficiently large" very soon); and the *quiescent phase*, which lasts from the end of the exponential growth phase until the time of recovery. The purpose of defining these two genetic phases is that under the stochastic pure-birth model described in Supplement A and the assumption of neutral evolution (i.e. no mutations have selective advantage or disadvantage), the only *de novo* iSNVs that rise to detectable frequency must occur early on in the viral replication process. Mutation events that occur when the viral population is already very large negligibly impact the ultimate fraction of virions to exhibit said mutation—even though birth events continue to occur frequently throughout the quiescent phase.

Hence, in our model, we explicitly track the composition of the viral population within each host at two points in time: when the host is inoculated, and at the end of the exponential growth phase. This allows us to apply our iSNV frequency model from Supplement A to the composition of the viral population at the end of the exponential growth phase, and then apply the Jukes-Cantor model, the simplest model of neutral evolution, to the change of this composition over the quiescent phase. Suppose that the exponential growth phase ends when the viral population reaches $k$ particles. How do we choose a reasonable value of $k$? Recall from Supplement A that our approximation of the PDF of $X$ relies on the idea that $p$, the probability that a birth event introduces a mutation, is small relative to $1/k$. However, we also want $k$ to be large enough such that any stochastic shocks to the composition over the viral population over the quiescent phase are negligible. In our model, we choose $k = \lfloor 1/\sqrt{p} \rfloor$ as a way to satisfy these two conditions; however, a wide range of other choices (e.g. $k = \lfloor p^{-2/3} \rfloor$ as in Specht and Mitzenmacher (2023)), are possible. For a formal analysis of the variance in the composition of the viral population over the quiescent phase, and for error bounds on the approximate distribution of mutated particles at the end of the exponential growth phase, see Specht and Mitzenmacher (2023).

#### B.2 Parameters and Data

With the theoretical motivation in place, we are now ready to begin defining our model. Let $N$ be the number of cases in the outbreak. Let $L$ be the length of the viral genome. Let $\boldsymbol{\theta}$ be a vector of unknown parameters—concretely,

$$\boldsymbol{\theta} = (\mu, p, w, \epsilon, \gamma, \alpha, \nu, \xi, \tau_E, \tau_T, \tau_I, \sigma_E, \sigma_T, \sigma_I, \mathbf{t}_E, \mathbf{t}_I, \mathbf{a}, \mathbf{h}, \mathbf{B}, \mathbf{X}),$$

where

- $\mu$ is the evolution rate of the virus, in substitutions per site per day.
- $p$ is probability of a mutation per site per cycle.
- $w$ is the number of virions produced per cycle.
- $\epsilon$ is the sequencing error rate, i.e., the probability that a nucleotide is misread as a different nucleotide.
- $\gamma + 1$ is the mean size of the transmission bottleneck, in number of virions (as any transmission bottleneck must have at least one virion).
- $\alpha + 1$ is the expected number of index cases (as any outbreak must have at least one index case).
- $\nu$ is the probability that a true transmission link appears in contact tracing data. If no contact tracing data is provided, this parameter is not used.
- $\xi$ is the probability that a pair of people who did *not* transmit to each other appears in contact tracing data. If no contact tracing data is provided, this parameter is not used.

- $\tau_E$ is the mean amount of time between the end of the exponential growth phase and the beginning of the infectious phase. (We assume the exponential growth phase always ends before the infectious phase begins).
- $\tau_T$ is the mean duration of the sojourn period (in days), defined as the interval between entering the infectious phase and receiving a diagnostic test.
- $\tau_I$ is the mean duration of the infectious period, in days.
- $\sigma_E^2$ is the variance in the amount of time between the end of the exponential growth phase and the beginning of the infectious phase, in days.
- $\sigma_T^2$ is the variance of the duration of the sojourn period, in days.
- $\sigma_I^2$ is the variance of the duration of the infectious period, in days.
- $\mathbf{t}_E$ is a vector of length $N$, whose $i$th entry $t_{Ei}$ is the time of exposure for case $i$.
- $\mathbf{t}_I$ is a vector of length $N$, whose $i$th entry $t_{Ii}$ is the time of the beginning of the infectious period for case $i$.
- $\mathbf{a}$ is a vector of length $N$, whose $i$th entry $a_i$ is the number of virions with which host $i$ was initially infected.
- $\mathbf{h}$ is a vector of length $N$, whose $i$th entry $h_i$ is the index of the infector of host $i$. If $i$ is an index case, we set $h_i = 0$.
- $\mathbf{B}$ is an $N \times L \times 4$ array, whose $(i, j, s)$th entry $b_{ijs}$ is the number of particles to inoculate host $i$ which exhibit allele $s$ at site $j$ on the viral genome. The values $s \in \{1, 2, 3, 4\}$ represent nucleotides {`A`, `C`, `G`, `T`}, respectively. We assume that at most two of $\{b_{ij1}, b_{ij2}, b_{ij3}, b_{ij4}\}$ are nonzero. Note that $b_{ij1} + b_{ij2} + b_{ij3} + b_{ij4} = a_i$ for all $j$. For ease of notation, let $\mathbf{b}_{ij} = (b_{ij1}, b_{ij2}, b_{ij3}, b_{ij4})$.
- $\mathbf{X}$ is an $N \times L \times 4$ array, whose $(i, j, s)$th entry $x_{ijs}$ is the fraction of the viral population to exhibit allele $s$ at site $j$ at the end of the exponential growth phase in host $i$. We assume that at most two of $\{x_{ij1}, x_{ij2}, x_{ij3}, x_{ij4}\}$ are nonzero. Note that $x_{ij1} + x_{ij2} + x_{ij3} + x_{ij4} = 1$ for all $i, j$. For ease of notation, let $\mathbf{x}_{ij} = (x_{ij1}, x_{ij2}, x_{ij3}, x_{ij4})$.

Next, let $\mathbf{Y}$ be a vector of known parameters—concretely,

$$\mathbf{Y} = (\mathbf{t}_T, \mathbf{R}, S)$$

where

- $\mathbf{t}_T$ is a vector of length $N$, whose $i$th entry $t_{Ti}$ is the time of testing for case $i$.
- $\mathbf{R}$ is an $N \times L \times 4$ array, whose $(i, j, s)$th entry $r_{ijs}$ is the number of reads exhibiting nucleotide $s$ at site $j$ on the viral genome of host $i$. For ease of notation, let $\mathbf{r}_{ij} = (r_{ij1}, r_{ij2}, r_{ij3}, r_{ij4})$.
- $S$ is a set of unordered pairs, where each pair represents a traced contact. Each unordered pair consists of two distinct integers ranging from 1 to $N$, inclusive.

Finally, we also define several variables which are deterministic functions of the known or unknown parameters, for notational convenience:

- $\mathbf{D}$ is an $N \times L$ matrix, whose $(i, j)$th entry $d_{ij}$ is the read depth at site $j$ in host $i$, equal to $r_{ij1} + r_{ij2} + r_{ij3} + r_{ij4}$.
- $k = \lfloor 1/\sqrt{p} \rfloor$, as in Section B.1.
- $\lambda = \frac{\mu}{p}$ is the exponential growth rate of the viral population, in birth events (cycles) per day. Specifically, the number of virions $t$ days after a host contracts the virus is $a_i w^{\lambda t}$, since each birth event produces $w$ virions.
- $\mathbf{t}_G$ is a vector of length $N$, whose $i$th entry $t_{Gi}$ is the time at which the exponential growth phase of host $i$ ends. Solving for $t$ in the definition of $\lambda$, we obtain

$$t_{Gi} = t_{Ei} + \frac{\log(k) - \log(a_i)}{\lambda \log(w)}.$$

Let $\pi(\boldsymbol{\theta}|\mathbf{Y})$ represent the posterior distribution of the unknown parameters $\boldsymbol{\theta}$; let $L(\boldsymbol{\theta}; \mathbf{Y})$ the likelihood function, and $\pi(\boldsymbol{\theta})$ the prior on $\boldsymbol{\theta}$. Our goal is to develop a suitable prior and likelihood function, which will allow us to approximate the posterior via Bayes' Theorem:

$$\pi(\boldsymbol{\theta}|\mathbf{Y}) \propto L(\boldsymbol{\theta}; \mathbf{Y}) \times \pi(\boldsymbol{\theta}).$$

##### B.3 Prior

Under the prior distribution, we assume all entries of the vector $\boldsymbol{\theta}$ to be independent, with the exception of $\mathbf{t}_E$, $\mathbf{t}_I$, and $\mathbf{a}$, whose prior distributions depend on other parameters in $\boldsymbol{\theta}$. Adopting the method of Lau et al. (2015), we propose nearly-uniform priors on $\mu, p, \epsilon, \gamma, \tau_E, \tau_T, \tau_I, \sigma_E^2, \sigma_T^2$, and $\sigma_I^2$:

- $\mu \sim$ Expo(0.001), restricted to the support $[0, 0.01]$.
- $p \sim$ Expo(0.001), restricted to the support $[0, 0.01]$.
- $\epsilon \sim$ Expo(0.001), restricted to the support $[0, 0.01]$.
- $\gamma \sim$ Expo(0.001), restricted to the support $[0, 10]$.
- $\tau_E \sim$ Expo(0.001), restricted to the support $[0, 10]$.
- $\tau_T \sim$ Expo(0.001), restricted to the support $[0, 10]$.
- $\tau_I \sim$ Expo(0.001), restricted to the support $[0, 10]$.
- $\sigma_E^2 \sim$ Expo(0.001), restricted to the support $[0.1, 15]$.
- $\sigma_T^2 \sim$ Expo(0.001), restricted to the support $[0.1, 15]$.
- $\sigma_I^2 \sim$ Expo(0.001), restricted to the support $[0.1, 15]$.

Once again following Lau et al., we impose a strong prior on $\alpha$, reflecting that among relatively self-contained outbreaks with robust testing (the main focus of our tool), being infected by an outside (unobserved) source is much rarer than being infected by an inside (observed) one:

- $\alpha \sim \text{Beta}(1, 1 \times 10^{10})$.

For $\nu$, $\xi$, $w$, $\mathbf{a}$, $\mathbf{h}$, and $\mathbf{X}$, we propose uniform priors:

- $\nu \sim \text{Unif}(0, 1)$.
- $\xi \sim \text{Unif}(0, 1)$.
- $w \sim \text{Unif}(2, 10^4)$.
- $h_i \sim \text{Unif}\{0, 1, \ldots, N\}$, i.i.d. for all $i$.
- $\mathbf{x}_{ij} \sim \text{Dirichlet}(1, 1, 1, 1)$, i.i.d. for all $i$ and $j$. Intuitively, this means that all length-4 vectors whose entries sum to 1 are equally probable.

Finally, we define the priors for the remaining parameters hierarchically, or in terms of the (fixed) data:

- $t_{Ti} - t_{Ii} \sim \text{Unif}(0, 20)$, i.i.d. for all $i$.
- $t_{Ii} - t_{Ei} | \mathbf{t}_I \sim \text{Unif}(0, 20)$, conditionally i.i.d. given $\mathbf{t}_I$ for all $i$.
- $a_i | \gamma \sim \text{Pois}(\gamma)$, conditionally i.i.d. given $\gamma$ for all $i$. For practical reasons, in the algorithmic implementation we set an upper limit to $a_i$ of 300 particles.
- $\mathbf{b}_{ij} | a_i$ is uniformly distributed over all length-4 vectors of non-negative integers with at most two nonzero entries, and whose entries sum to $a_i$. We model $\mathbf{b}_{ij} | a_i$ conditionally i.i.d. across all $j$ given $a_i$, and independent across $i$.

##### B.4 Likelihood

Before proceeding, we define a few additional functions to help us express the likelihood in an interpretable form:

- $f_{\text{Beta}}(x; \alpha, \beta)$ and $F_{\text{Beta}}(x; \alpha, \beta)$ are the PDF and CDF of the Beta Distribution evaluated at $x$ with parameters $\alpha$ and $\beta$, respectively.
- $f_{\text{Gamma}}(x; \alpha, \lambda)$ and $F_{\text{Gamma}}(x; \alpha, \lambda)$ are the PDF and CDF of the Gamma Distribution evaluated at $x$ with shape parameters $\alpha$ rate parameter $\lambda$, respectively.
- $f_{\text{Pois}}(x; \lambda)$ and $F_{\text{Pois}}(x; \lambda)$ are the PMF and CDF of the Poisson Distribution evaluated at $x$ with parameters $\lambda$, respectively.
- $f_{\text{Bin}}(x; N, p)$ and $F_{\text{Bin}}(x; N, p)$ are the PMF and CDF of the Binomial Distribution evaluated at $x$ with size parameter $N$ and probability parameter $p$, respectively.
- $f_{\text{Mult}_k}(x; N, \mathbf{p})$ and $F_{\text{Mult}_k}(\mathbf{x}; N, \mathbf{p})$ are the PMF and CDF of the $k$-dimensional Multinomial Distribution evaluated at $\mathbf{x}$ with size parameter $N$ and probability vector $\mathbf{p}$, respectively.

Let us now define the likelihood function. We express the function as the product of two terms:

$$L(\boldsymbol{\theta}; \mathbf{Y}) = L_e(\boldsymbol{\theta}; \mathbf{Y}) \times L_g(\boldsymbol{\theta}; \mathbf{Y})$$

where $L_e$ is the likelihood of the epidemiological data and $L_g$ is the likelihood of the genetic data. The epidemiological likelihood function reflects a standard stochastic-epidemic process, with additional terms to account for the false-positive and false-negative reporting rate for contact tracing data, if provided:

$$L_e(\boldsymbol{\theta}; \mathbf{Y}) = \prod_{i \leq N} f_{\text{Gamma}}(t_{Ti} - t_{Ii}; \tau_T^2 / \sigma_T^2, \tau_T / \sigma_T^2) \tag{B.1}$$

$$\times \prod_{i \le N} f_{\text{Gamma}}\left(t_{Ii} - t_{Gi}; \tau_E^2/\sigma_E^2, \tau_E/\sigma_E^2\right) \tag{B.2}$$

$$\times \prod_{\substack{i \le N \\ h_i \neq 0}} (1 - F_{\text{Gamma}}(t_{Ei} - t_{Ih_i}; \tau_I^2/\sigma_I^2, \tau_I/\sigma_I^2)) \tag{B.3}$$

$$\times f_{\text{Bin}}\left(|\{i : h_i = 0\}| - 1; N - 1, \alpha\right) \tag{B.4}$$

$$\times \prod_{\substack{i \le N \\ h_i \neq 0}} \nu^{\mathbb{1}_{\{h_i, i\} \in S}} (1 - \nu)^{\mathbb{1}_{\{h_i, i\} \notin S}} \tag{B.5}$$

$$\times \prod_{\substack{1 \le i < j \le n \\ h_i \neq j \\ h_j \neq i}} \xi^{\mathbb{1}_{\{i,j\} \in S}} (1 - \xi)^{\mathbb{1}_{\{i,j\} \notin S}} \tag{B.6}$$

Note that the term B.3 computes the probability that each host's infector is in the infectious compartment at the time of transmission, and term B.4 computes the likelihood associated with the number of index cases. In terms B.5 and B.6, $\mathbb{1}_A$ denotes the indicator function of an event $A$.

We construct $L_g$ in several steps. The first step is to quantify the distribution of $\mathbf{x}_{ij}$, the composition of the viral population at site $j$ at the end of the exponential growth phase in host $i$, as a function of $\mathbf{b}_{ij}$, the contents of the inoculum at site $j$ in host $i$. Let $\rho(\mathbf{x}_{ij}; \mathbf{b}_{ij})$ represent the conditional probability density function of $\mathbf{x}_{ij}$ given $\mathbf{b}_{ij}$. Recall that we assume at most two of the entries of $\mathbf{b}_{ij}$ may be nonzero. In the case that *exactly* two are nonzero, let $s_1 < s_2$ denote the two nonzero entries of $\mathbf{b}_{ij}$. As in Supplement A, we define

$$\rho(\mathbf{x}_{ij}; \mathbf{b}_{ij}) = \begin{cases} f_{\text{Beta}}(x_{ijs_1}; b_{ijs_1}, b_{ijs_2}), & x_{ijs_1} + x_{ijs_2} = 1 \text{ and the other two entries of } \mathbf{x}_{ij} \text{ are } 0 \\ 0, & \text{otherwise} \end{cases}.$$

And, when exactly one entry, $s$, of $\mathbf{b}_{ij}$ is nonzero, we invoke the definition of $g$ in Supplement A:

$$\rho(\mathbf{x}_{ij}; \mathbf{b}_{ij}) = \begin{cases} g_{k,0,b_{ijs}}(1 - x_{ijs}), & \text{at most one } \mathbf{x}_{ijs'} \text{ is nonzero for } s' \neq s, \text{ and } \sum_{s \le 4} x_{ijs} = 1 \\ 0, & \text{otherwise} \end{cases}.$$

Now that we have defined the probability distribution for the composition of the viral population at the end of the exponential growth phase, let us quantify the genetic evolution that occurs over the course of the quiescent phase. We adopt the Jukes-Cantor model for this purpose, which states that the composition of the viral population changes approximately linearly over time with evolution rate $\mu$, and that the probabilities of any nucleotide being substituted for any other nuclotide are equal. Define a matrix $\mathbf{P}_{\mu,t}$ by

$$\mathbf{P}_{\mu,t} = \begin{pmatrix} 1 - \mu t & \frac{\mu t}{3} & \frac{\mu t}{3} & \frac{\mu t}{3} \\ \frac{\mu t}{3} & 1 - \mu t & \frac{\mu t}{3} & \frac{\mu t}{3} \\ \frac{\mu t}{3} & \frac{\mu t}{3} & 1 - \mu t & \frac{\mu t}{3} \\ \frac{\mu t}{3} & \frac{\mu t}{3} & \frac{\mu t}{3} & 1 - \mu t \end{pmatrix}.$$

Then, for $\mu t << 1$, the composition of the viral population in host $i$ at site $j$ is $\mathbf{P}_{\mu,t} \cdot \mathbf{x}_{ij}$, $t$ days after the end of the exponential growth phase.

We quantify sequencing error in similar fashion, by assuming all possible misreadings of one nucleotide as a different nucleotide are equally likely. Define a matrix $\mathbf{E}_\epsilon$ by

$$\mathbf{E}_\epsilon = \begin{pmatrix} 1-\epsilon & \frac{\epsilon}{3} & \frac{\epsilon}{3} & \frac{\epsilon}{3} \\ \frac{\epsilon}{3} & 1-\epsilon & \frac{\epsilon}{3} & \frac{\epsilon}{3} \\ \frac{\epsilon}{3} & \frac{\epsilon}{3} & 1-\epsilon & \frac{\epsilon}{3} \\ \frac{\epsilon}{3} & \frac{\epsilon}{3} & \frac{\epsilon}{3} & 1-\epsilon \end{pmatrix}.$$

Recall that the composition of the viral population at site $j$ in host $i$ at time $t_{Ti}$, conditional on $\mathbf{x}_{ij}$, is given by $\mathbf{P}_{\mu,t_{Ti}-t_{Gi}} \cdot \mathbf{x}_{ij}$. Hence, the probabilities of the sequencing machine reading each possible nucleotide at $j$ are given by $\mathbf{E}_\epsilon \cdot \mathbf{P}_{\mu,t_{Ti}-t_{Gi}} \cdot \mathbf{x}_{ij}$. Thus, we model sequencing read data by assuming that each read of site $j$ in host $i$ is an independent draw from the categorical distribution on the set of nucelotides $\{\mathtt{A}, \mathtt{C}, \mathtt{G}, \mathtt{T}\}$, with probabilities $\mathbf{E}_\epsilon \cdot \mathbf{P}_{\mu,t_{Ti}-t_{Gi}} \cdot \mathbf{x}_{ij}$, i.e.:

$$\mathbf{r}_{ij} | \mathbf{x}_{ij} \sim \mathrm{Mult}_4(d_{ij}, \mathbf{E}_\epsilon \cdot \mathbf{P}_{\mu,t_{Ti}-t_{Gi}} \cdot \mathbf{x}_{ij})$$

In the same way, we model the viral particles the inoculate host $i$ as independent categorical draws from the viral population of the *ancestor* of host $i$, as long as $i$ is not an index case:

$$\mathbf{b}_{ij} | \mathbf{x}_{h_i j} \sim \mathrm{Mult}_4(a_i, \mathbf{P}_{\mu,t_{Ei}-t_{Gh_i}} \cdot \mathbf{x}_{h_i j}), \quad \text{when } i \text{ is not an index case.}$$

Finally, we must consider the scenario in which $i$ is an index case. To assign a likelihood to the inoculum of an index case $i$, we ask the question: what is the probability that an unobserved case in the general population exhibits nucleotide $s$ at site $j$ on the viral genome? We model this probability as the fraction of reads of site $j$ to exhibit nucleotide $s$, *averaged across the entire observed outbreak.* Concretely, define an $L \times 4$ matrix $\mathbf{Q} = (q_{js})$ by

$$q_{js} = \frac{\sum_{i \le N} r_{ijs}}{\sum_{i \le N} d_{ij}}.$$

Then we interpret $q_{js}$ as the probability that an unobserved case $u$ exhibits nucleotide $s$ at site $j$ at the end of the exponential growth phase in $u$. (We assume that no iSNVs arise in the exponential growth phase of $u$). Let $\mathbf{1}_s$ denote the vector of length 4 whose $s$th entry equals 1, and whose other entries equal 0. Assuming that the interval between the end of the exponential growth phase in $u$ and the time at which $u$ transmits to index case $i$ is always a fixed 3 days (as we have no data to assume otherwise, we compute the probability density of the inoculum of host $i$ by averaging over the possible genetic composition of site $j$ in host $u$:

$$\mathbf{b}_{ij} \sim \sum_{s \le 4} \big( q_{js} \times \mathrm{Mult}_4(a_i, \mathbf{P}_{\mu,3} \cdot \mathbf{1}_s) \big), \quad \text{when } i \text{ is an index case.}$$

And finally, the probability that inoculum infecting host $i$ consists of $a_i$ virions is given by $f_{\mathrm{Pois}}(a_i - 1; \gamma)$. We are now ready to write the genomic likelihood function:

$$\begin{aligned} L_g(\boldsymbol{\theta}; \mathbf{Y}) = & \prod_{i \le N} \prod_{j \le L} \rho(\mathbf{x}_{ij}; \mathbf{b}_{ij}) \\ & \times \prod_{i \le N} \prod_{j \le L} f_{\mathrm{Mult}_4}\left(\mathbf{r}_{ij}; d_{ij}, \mathbf{E}_\epsilon \cdot \mathbf{P}_{\mu,t_{Ti}-t_{Gi}} \cdot \mathbf{x}_{ij}\right) \\ & \times \prod_{\substack{i \le N \\ h_i \neq 0}} \prod_{j \le L} f_{\mathrm{Mult}_4}(\mathbf{b}_{ij}; a_i, \mathbf{P}_{\mu,t_{Ei}-t_{Gh_i}} \cdot \mathbf{x}_{h_i j}) \end{aligned}$$

$$\times \prod_{\substack{i\le N\\ h_i\neq 0}} \prod_{j\le L} \sum_{s\le 4} q_{js} f_{\text{Mult}_4}\left(\mathbf{b}_{ij}; a_i, \mathbf{P}_{\mu,3}\cdot \mathbf{1}_s\right)$$

$$\times \prod_{i\le N} f_{\text{Pois}}(a_i - 1; \gamma).$$

##### B.5 Applying a Limit of Detection Filter

The expressions we derived for $L_e(\boldsymbol{\theta}; \mathbf{Y})$ and $L_g(\boldsymbol{\theta}; \mathbf{Y})$ theoretically quantify the likelihood of any viral outbreak with deep-sequencing. In practice, however, it may be useful to apply a limit of detection filter for sequencing data, since iSNV calls below a certain frequency $U$ (often 3%) or with fewer than some number $M$ of alternate allele reads (often 10) are usually considered unreliable. To account for this, we revise the conditional probability distribution of $\mathbf{r}_{ij}|\mathbf{x}_{ij}$ by summing over all possible vectors $\mathbf{r}'_{ij}$ that would equal $\mathbf{r}_{ij}$ after applying these two filters. Concretely, define a filter function $z : \mathbb{N}^4 \to \mathbb{N}^4$ by

$$z(\mathbf{r}_{ij})_s \begin{cases} 0, & r_{ijs} < \max(d_{ij}U, M) \\ r_{ijs}, & \max(d_{ij}U, M) \le r_{ijs} \le \min(d_{ij}(1-U), d_{ij} - M) \\ d_{ij}, & r_{ijs} > \min(d_{ij}(1-U), d_{ij} - M) \end{cases}.$$

Then we redefine the conditional PMF of $\mathbf{r}_{ij}|\mathbf{x}_{ij}$ as:

$$\sum_{\{\mathbf{r}'_{ij}: z(\mathbf{r}'_{ij}) = \mathbf{r}_{ij}\}} f_{\text{Mult}_4}(\mathbf{r}'_{ij}; d_{ij}, \mathbf{E}_\epsilon \cdot \mathbf{P}_{\mu, t_{Ti} - t_{Gi}} \cdot \mathbf{x}_{ij}).$$

### C MCMC Implementation

We sampled the posterior distribution of our outbreak reconstruction model using the Metropolis-Hastings Algorithm for Markov Chain Monte Carlo (MCMC). We ran each chain for 10,000 MCMC iterations with a burnin of 10%. Here, we describe the initialization and execution of the algorithm in greater detail.

##### C.1 Initialization

We initialized the Markov chain with the following parameter values:

- $\mu = 1 \times 10^{-6}$.
- $p = 1 \times 10^{-6}$.
- $w = 1000$.
- $\epsilon = 1 \times 10^{-10}$.
- $\gamma = 2$.
- $\alpha = 1 \times 10^{-10}$.
- $\nu = 0.5$.
- $\xi = 0.5$.

- $\tau_E = 2$.
- $\tau_T = 2$.
- $\tau_I = 6$.
- $\sigma_E^2 = 2$.
- $\sigma_T^2 = 2$.
- $\sigma_I^2 = 6$.
- $t_{Ii} = t_{Ti} - 2$, for all $i$.
- $t_{Ei} = t_{Ii} - 2$, for all $i$.
- $a_i = 1$, for all $i$.
- $h_i = 0$, for all $i$, i.e. everyone starts out as an index case.
- $x_{ijs} = \frac{r_{ijs}}{d_{ij}}$, rounded to the nearest integer.
- **B** is listed as a parameter of our model for ease of explanation; however, integrating it out still yields a closed analytic form of the likelihood function.

#### C.2 Moveset

To execute the Metropolis-Hastings algorithm, we employed the following moveset:

- Adjust $\mu$ by $\mathcal{N}(0, 1 \times 10^{-6})$ nt site$^{-1}$ day$^{-1}$.
- Adjust $p$ by $\mathcal{N}(0, 1 \times 10^{-6})$ nt site$^{-1}$ cycle$^{-1}$.
- Adjust $w$ by $\mathcal{N}(0, 50)$ virions.
- Adjust $\epsilon$ by $\mathcal{N}(0, 1 \times 10^{-12})$ units.
- Adjust $\gamma$ by $\mathcal{N}(0, 0.5)$ units.
- Adjust $\alpha$ by $\mathcal{N}(0, 1 \times 10^{-9})$ units.
- Adjust $\nu$ by $\mathcal{N}(0, 0.1)$ units.
- Adjust $\xi$ by $\mathcal{N}(0, 0.1)$ units.
- Adjust $\tau_E$ by $\mathcal{N}(0, 0.5)$ days.
- Adjust $\tau_T$ by $\mathcal{N}(0, 0.5)$ days.
- Adjust $\tau_I$ by $\mathcal{N}(0, 0.5)$ days.
- Adjust $\sigma_E^2$ by $\mathcal{N}(0, 0.5)$ days$^2$.
- Adjust $\sigma_T^2$ by $\mathcal{N}(0, 0.5)$ days$^2$.
- Adjust $\sigma_I^2$ by $\mathcal{N}(0, 0.5)$ days$^2$.

- Adjust $x_{ij}$ by $\mathcal{N}(0, 0.01)$ units.
- With probability 1/2, increase $a_i$ by 1, and with probability 1/2, decrease $a_i$ by 1 (unless $a_i = 1$, in which case, do nothing).
- Set $t_{Ei}$ to a uniform random value between $t_{Ih_i}$ and $t_{Ei}$.
- Set $t_{Ii}$ to a uniform random value between $t_{Ei}$ and the minimum over $t_{Ti}$ and the times at which case $i$ transmits to other cases.
- Set $h_i$ to any host who becomes infectious before $t_{Ei}$.
- Set $h_i$ to any host who becomes infectious before $t_{Ei}$. Let $h'_i$ denote the new ancestor of host $i$. Then, swap $x_{h_i j}$ with $x_{h'_i j}$ at all sites $j$ for which $|x_{h_i j} - x_{h'_i j}| > 1/2$.
- Set $t_{Ei}$ and $t_{Ii}$ to the minimum and maximum of two uniform random draws between $t_{Ih_i}$ and the minimum over $t_{Ti}$ and the times at which case $i$ transmits to other cases. Let $t'_{Ei}$ denote the proposed new value of $t_{Ei}$. Then, set $h_i$ to any host who becomes infectious before $t'_{Ei}$.
- Set $h_i$ to any host who becomes infectious before $t_{Ei}$. Let $h'_i$ be the proposed new value of $h_i$, i.e. the proposed new ancestor of host $i$. Then, set $t_{Ei}$ to a uniform random value between $t_{Ih'_i}$ and $t_{Ei}$.
- Identify a transmission chain of three hosts, i.e. some hosts $f, g, i$ such that $f$ transmits to $g$ and $g$ transmits to $i$. Rearrange this transmission chain such that $f$ transmits to $i$ and $i$ transmits to $g$. Also, swap $t_{Eg}$ and $t_{Ei}$, $t_{Ig}$ and $t_{Ii}$, and $a_g$ and $a_i$.
- Perform the above move. Then, also swap the children of hosts $g$ and $i$, i.e. reassign the ancestor of anyone infected by host $i$ to be host $g$, and vice versa.

#### C.3 Approximations

For computational efficiency, we apply two approximations. First, for all sites $j$ where every read across the entire outbreak exhibits the same allele (i.e. sites $j$ for which at most one of $q_{j1}, q_{j2}, q_{j3}, q_{j4}$ is nonzero), we do not store values of $x_{ij}$ for any $i$. Instead, we approximate the probability of observing no iSNV at site $j$ in host $i$ as

$$(1-p)^k + (1-(1-p)^k)G_k(\min(L, d_{ij}M)),$$

i.e. the probability that the minor allele fraction at the end of the exponential growth phase does not exceed the limit of detection. (See Supplement A for the definition of $G_k$).

For the other approximation, in practice, we chose to leave $\sigma_E, \sigma_T, \sigma_I$, and $\tau_I$ fixed (ignoring the aforementioned MCMC moves for these parameters). We observed that $\mathbf{t}_E$ and $\mathbf{t}_I$ were overwhelmingly influenced by the transmission network structure and genomic parameters, and minimally by $\sigma_E, \sigma_T$. While these approximations mean our model does not infer $\sigma_E, \sigma_T, \sigma_I$, and $\tau_I$, inferences of other parameters remained largely unchanged.
